# Geographic Disparities and Determinants of COVID-19 Incidence Risk in the Greater St. Louis Area, Missouri

**DOI:** 10.1101/2021.12.18.21268027

**Authors:** Praachi Das, Morganne Igoe, Suzanne Lenhart, Lan Luong, Cristina Lanzas, Alun L. Lloyd, Agricola Odoi

## Abstract

**Background:** Evidence suggests that the risk of Coronavirus Disease 2019 (COVID-19) varies geographically due to differences in population characteristics. Therefore, the objectives of this study were to identify: (a) geographic disparities of COVID-19 risk in the Greater St. Louis area of Missouri, USA; (b) predictors of the identified disparities.

**Methods:** Data on COVID-19 incidence and chronic disease hospitalizations were obtained from the Departments of Health and Missouri Hospital Association, respectively. Socioeconomic and demographic data were obtained from the 2018 American Community Survey while population mobility data were obtained from the SafeGraph website. Choropleth maps were used to identify geographic disparities of COVID-19 risk and its predictors at the ZIP Code Tabulation Area (ZCTA) spatial scale. Global negative binomial and local geographically weighted negative binomial models were used to identify predictors of ZCTA-level geographic disparities of COVID-19 risk.

**Results:** There were geographic disparities in COVID-19 risk. Risks tended to be higher in ZCTAs with high percentages of the population with a bachelor’s degree (p<0.0001) and obesity hospitalizations (p<0.0001). Conversely, risks tended to be lower in ZCTAs with high percentages of the population working in agriculture (p<0.0001). However, the association between agricultural occupation and COVID-19 risk was modified by per capita between ZCTA visits. Areas that had both high per capita between ZCTA visits and high percentages of the population employed in agriculture had high COVID-19 risks. The strength of association between agricultural occupation and COVID-19 risk varied by geographic location.

**Conclusions:** Geographic Information Systems, global and local models are useful for identifying geographic disparities and predictors of COVID-19 risk. Geographic disparities of COVID-19 risk exist in the St. Louis area and are explained by differences in sociodemographic factors, population movements, and obesity hospitalization risks. The latter is particularly concerning due to the growing prevalence of obesity and the known immunological impairments among obese individuals. Therefore, future studies need to focus on improving our understanding of the relationships between COVID-19 vaccination efficacy, obesity and waning of immunity among obese individuals so as to better guide vaccination regimens, reduce disparities and improve population health for all.

## Background

Since the first reported case in Wuhan (China) in December 2019, Coronavirus Disease 2019 (COVID-19) has spread worldwide with over 273 million confirmed cases and over 5.3 million deaths as of December 18, 2021 [1]. Within the United States, there have been over 50 million confirmed cases and over 805,000 reported deaths with Missouri reporting over 961,000 confirmed cases and 15,000 deaths [2,3].

There is evidence of geographic disparities in incidence risk of COVID-19 and that these disparities are determined, at least in part, by population characteristics and movement behavior that if identified may guide disease control. For instance, some studies have reported higher prevalence of COVID-19 in regions with high densities of Black and/or Hispanic populations [4,5]. There have also been reports of associations between COVID-19 incidence risk and both healthcare and non-healthcare related occupations [6–8]. During the initial stages of the COVID-19 pandemic, stay-at-home and social distancing measures were implemented to minimize contacts between infected or potentially infected and susceptible individuals so as to curb disease transmission [9,10]. These measures were based on the understanding that more interactions between infected and susceptible individuals in the population would lead to more cases, reflecting the potential association between COVID-19 incidence risk and the movement of individuals. Chang and co-workers used population mobility, derived from SafeGraph mobile phone data, and susceptible-exposed-infectious-removed (SEIR) model and predicted high infection rates among disadvantaged racial and socioeconomic groups [11]. The study noted that the identified differences were solely due to differences in mobility since disadvantaged groups were not able to reduce their mobility as much as their non-disadvantaged counterparts and that they tended to visit crowded places that were inherently associated with higher infection/disease risks [11].

Understanding the geographic disparities in the distribution of COVID-19 risk and identifying the determinants/predictors of these disparities at the local level is critically important in identifying communities at highest risk. This information is important in guiding local health policies as well as resource allocation for targeting interventions such as distribution of testing kits and vaccines. Thus, the objective of this study was to identify geographic disparities in COVID-19 incidence risk in the greater St. Louis area of Missouri and to investigate sociodemographic, chronic condition and population movement determinants/predictors of the identified disparities.

## Methods

### Study Area

This retrospective ecological study was conducted in Missouri in an area that included a total of 108 ZIP Code Tabulation Areas (ZCTAs) located in 6 counties: Franklin, Jefferson, St. Charles, St. Louis City, St. Louis, and Warren counties (**Figure 1**). The area has a total population of approximately 2 million people comprised of about 20% Black, 3% Hispanic or Latino, 3% Asian and 74% White individuals. Forty-eight percent of the population was male, while 52% was female. The ZCTA-level population density ranged from 10 people per square mile in St. Charles County to 9,368 in St. Louis City County.

**Figure 1.**
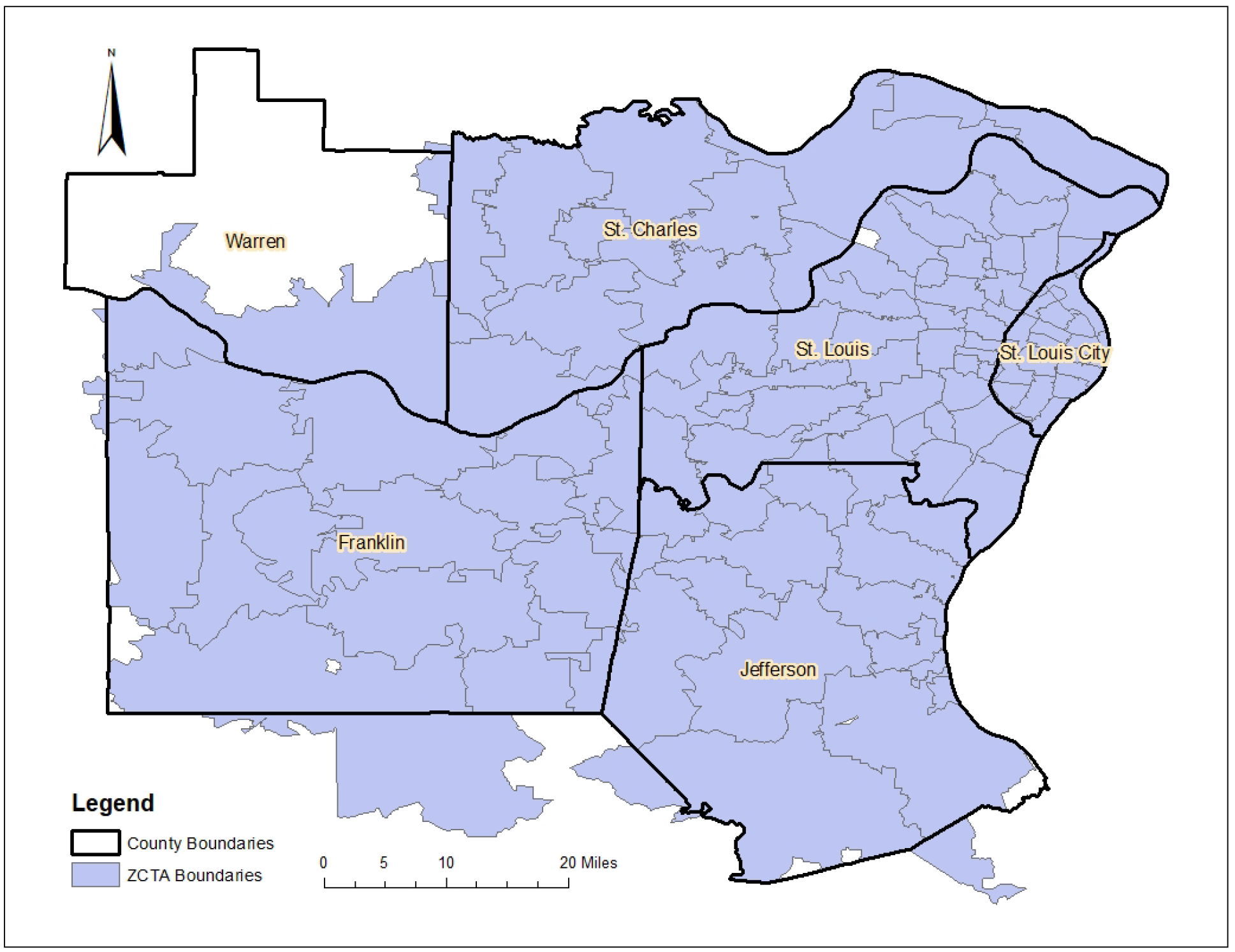
Map of study the area showing geographic distribution of ZCTAs and Counties of the Greater St. Louis Area, Missouri (USA)

### Data Sources

#### Confirmed COVID-19 Data

Data on confirmed cases of COVID-19 reported in the study area between March 23^rd^ and October 8^th^, 2020 were obtained from the county Departments of Health. These data were aggregated to the ZCTA-level to obtain cumulative incidence risks of confirmed COVID-19 cases over the study period for each ZCTA.

#### Sociodemographic and Base Map Data

Data on ZCTA-level sociodemographic factors, including age, sex, racial composition, population density, household size, education level, employment industry (occupation), and median income were obtained from the U.S. Census Bureau 2018 American Community Survey (ACS) 5-year estimates [12]. Cartographic boundary files used as base maps for all cartographic displays were obtained from the US Census Bureau Website [13].

#### Chronic Disease Data

Data on hospitalizations attributed to chronic conditions, by ZCTA of residence, were obtained from the Hospital Industry Data Institute, a not-for-profit organization founded by the Missouri Hospital Association that summarizes state-level hospital discharge data. The data included the number of unique patient hospitalizations with ICD-10 codes for obesity, tobacco use, cancer, chronic obstructive pulmonary disease (COPD), chronic kidney disease, heart failure, and diabetes. These variables were selected due to their hypothesized potential associations with COVID-19 risk. Therefore, risks of hospitalization with each of the above conditions were computed and used in subsequent analyses to investigate their associations with COVID-19 risk.

#### Movement Data

Data on daily movement patterns of individuals into and within the study area from March 23^rd^ to October 8^th^ 2020 were obtained from the Social Distancing Metrics cellphone dataset of SafeGraph [14]. SafeGraph is a company that aggregates anonymized location data from smartphones [11]. The data are useful for studying population mobility patterns. The metrics selected from the dataset were the origin census block group (CBG), which represents the home location of the owner of a device, and the destination CBG. For privacy reasons, the data did not include CBGs which had less than 5 recorded devices. To allow ZCTA-level analysis, point-in-polygon joins were used to link CBGs to ZCTAs using CBG geographic centroids and ZCTA polygon data obtained from the U.S. Census Bureau [13]. A total of 5 CBGs did not match any ZCTA and could not be linked to the ZCTA polygon file. Thus, the final dataset had a total of 103 of the original 108 ZCTAs in the study area. Therefore, all subsequent analyses included 103 ZCTAs.

Population movement metrics were defined to describe the different types of movements observed in the study area. Travel from outside the study area into one of the 103 study area ZCTAs was called an “***outside visit***”. Travel from one study area ZCTA to another study area ZCTA was referred to as a “***between visit***” while travel within a study area ZCTA was called a “***within visit***”. Each of these movement metrics was expressed either as: (a) *raw values*: total number of visits in a ZCTA or (b) *per capita (population-adjusted) values*: total number of visits in a ZCTA divided by the population of the ZCTA. The per capita values were particularly important in situations where there were disproportionate numbers of movements in a ZCTA relative to the size of the ZCTA’s population (e.g. a large number of visits to sparsely populated ZCTAs).

### Descriptive Analysis and identification of geographic disparities in COVID-19 Risk

Descriptive analyses were performed in R version 4.1.0 [17] and implemented in RStudio version 1.4.1103 [18]. The Shapiro-Wilk test was used to assess for normality of distribution of continuous variables. Medians as well as 1^st^ and 3^rd^ quartiles were reported for each variable since all were non-normally distributed (**Table 1**). To investigate geographic disparities in COVID-19 incidence risk, choropleth maps were generated in QGIS [16] using Jenk’s optimization classification scheme [17] to determine the break-points.

**Table 1:** Summary statistics of ZCTA-level potential predictors of COVID-19 risk in the Greater St. Louis Area, Missouri (USA)

| Type of Variable | Variable | Median | 1<sup>st</sup> Quartile | 3<sup>rd</sup> Quartile |
| --- | --- | --- | --- | --- |
| Demographic Factors |  |  |  |  |
|  | % male population | 48.6 | 47.4 | 50.3 |
|  | % black population | 3.7 | 0.9 | 34.2 |
|  | % Hispanic/Latino population | 2.2 | 1.0 | 3.2 |
|  | % over 65 | 15.0 | 12.3 | 17.8 |
|  | Average household size | 2.5 | 2.3 | 2.7 |
| Economic Variables |  |  |  |  |
|  | % below poverty level | 9.5 | 5.8 | 16.8 |
|  | median household income | 59,769 | 46,005 | 77,504 |
| Educational Variables |  |  |  |  |
| | % with ≤ high school education | 38.2 | 23.9 | 47.6 |
|  | % with some college education | 23.0 | 19.8 | 25.6 |
|  | % with associate's degree | 8.6 | 6.6 | 10.6 |
|  | % with bachelor's degree | 18.0 | 9.4 | 25.9 |
| Occupation Variables |  |  |  |  |
|  | % in agriculture<sup>1</sup> | 0.5 | 0.2 | 1.0 |
|  | % in construction | 5.2 | 3.38 | 9.5 |
|  | % in retail | 10.8 | 8.3 | 12.0 |
|  | % in transportation | 4.45 | 3.1 | 6.05 |
|  | % in manufacturing | 11.0 | 8.7 | 13.0 |
|  | % in education or healthcare | 23.6 | 20.6 | 27.7 |
| Health Behavior (ZCTA-level number of hospitalized patients that use tobacco per 100 persons) |  |  |  |  |
|  | % tobacco<sup>2</sup> | 10.4 | 6.8 | 14.5 |
| Co-morbidities (ZCTA-level number of hospitalized patients with specific condition per 100 persons) |  |  |  |  |
|  | % obesity | 7.0 | 5.6 | 8.0 |
|  | % cancer | 3.8 | 3.3 | 4.3 |
|  | % COPD<sup>3</sup> | 4.0 | 3.1 | 4.9 |
|  | % CKD<sup>4</sup> | 7.6 | 6.4 | 8.9 |
|  | % heart failure | 3.2 | 2.6 | 3.8 |
|  | % diabetes | 7.5 | 6.5 | 9.2 |
| Movement variables |  |  |  |  |
|  | Outside visits | 44,501 | 16,974 | 71,724 |
|  | Within visits | 181,031 | 78,776 | 401,433 |
|  | Between visits | 163,655 | 71,337 | 298,681 |
|  | Per capita outside visits | 2.4 | 1.5 | 4.3 |
|  | Per capita within visits | 13.2 | 9.0 | 18.8 |
|  | Per capita between visits | 9.7 | 6.8 | 1.4 |

<sup>1</sup>Percent of population employed in agriculture, forestry, fishery and mining
<sup>2</sup>ZCTA-level % hospitalized patients that were tobacco users
<sup>3</sup>Chronic Obstructive Pulmonary Disease;
<sup>4</sup>Chronic Kidney Disease

### Investigation of Predictors of the geographic disparities of COVID-19 Risk

#### Global Models

Univariable global Poisson models were used to investigate associations between each of the predictors and COVID-19 incidence risk. The response variable of interest was the ZCTA-level number of confirmed COVID-19 cases offset by the natural log of the population in each ZCTA. The predictors investigated were ZCTA-level sociodemographic factors, chronic condition risks, and movement patterns between and within ZCTAs. The models were fit using the Generalized Linear Model framework (glm function) in R [15], specifying Poisson distribution and log link. Variables with a p-value less than 0.15 were considered potentially associated with the outcome and investigated further. The linear relationships between the log of the incidence risk and the potential predictors were assessed graphically.

Bivariate Spearman rank correlation analyses were used to identify pairs of highly correlated (r≥0.7) potential predictors. To avoid multicollinearity during the subsequent multivariable modeling, only one of a pair of highly correlated variables were assessed in the multivariable model. Backwards elimination process was then used to build a multivariable Poisson model using an alpha of 0.05 to assess statistical significance. The final Poisson model showed evidence of overdispersion based on the fact that the ratio of residual deviance to residual degrees of freedom was much greater than 1, implying the Poisson model was not appropriate for the data.

To account for overdispersion, a negative binomial (NB) model was fit to the data using the same approach described above for Poisson model. The NB model was fit using the glm.nb function of the MASS package in R [15] specifying negative binomial distribution and log link. Potential confounding variables were investigated by assessing if removal of a suspected confounding variable from the model resulted in a change of at least 20% of the coefficients of any of the variables still in the model. If this happened, then the suspected confounding variable would be retained in the model regardless of its statistical significance, otherwise, it would be removed from the model unless it had a significant p-value in which case it would be retained in the model as an important predictor. Two-way biologically meaningful interaction terms between predictors in the final main effects multivariable model were assessed and significant ones retained in the model. Goodness-of-fit of the final multivariable negative binomial model was assessed using Pearson and Deviance chi-square goodness-of-fit tests in STATA 16.1 [18].

#### Local Models

A geographically weighted negative binomial (GWNB) model, proposed by Silva and Rodrigues [19], was fit to the data to assess if the association between each of the predictors and COVID incidence risks varied by geographic location. The same outcome, offset, distribution, link function, and predictors used in the final global multivariable NB model were used for this model. The GWNB theoretically estimates as many regression coefficients as the number of ZCTAs in the study area. This is unlike the global multivariable model which estimates one regression coefficient for each predictor and assumes that the strength of association with the outcome remains constant across all ZCTAs in the study area.

SAS/IML macro [20] of SAS version 9.4 [21] was used to fit the local GWNB model. The estimation of local regression coefficients was based on a biquadratic kernel weighting function [20], while the bandwidth was estimated using the adaptive method which allows the size of the bandwidth to vary based on the density of observations. Bias-corrected Akaike Information Criteria (AICc) was used to determine the optimum kernel bandwidth and compare the goodness-of-fit of the global NB and GWNB models. A lower AICc value indicated a better fitting model.

Stationarity of the GWNB coefficients were assessed using: (a) randomization non-stationarity test based on 999 replications [22]; (b) comparison of the interquartile range of the local GWNB model coefficients with the standard error estimates of the global NB model. Local coefficients with interquartile ranges larger than twice the standard error of the regression coefficient from the global NB model were considered non-stationary [23,24].

## Results

### Descriptive statistics

The median percentages of male, Black, and Hispanic populations across ZCTAs were 48.6%, 3.7% and 2.2% of the study population, respectively, while the median percentage of individuals aged ≥65 years was 15% (**Table 1**). The median average household size was 2.5 and the median household income was $59,769 with 9.5% of the population living below the poverty line. Regarding education, individuals with a high school or lower education comprised the highest percentage (38.2%) of the population followed by those with some college education (23%). As for occupation, most of the population worked in education or healthcare (23.6%), followed by manufacturing (11%) and the least were employed in agriculture (0.5%) (**Table 1**).

Of the chronic conditions assessed for potential association with COVID-19, chronic kidney disease (CKD) had the highest median hospitalization risk (7.6%) while heart failure had the lowest (3.2%). Regarding movement of the population, the largest median number of movements across all study ZCTAs were within ZCTA visits (median: 181,031) while the fewest number of visits were outside visits (median: 44,501). The values of the per capita (population adjusted) visits followed the same ordering as the above raw (unadjusted) visits with the highest median number of per capita visits being within ZCTA visits (median: 13.2) and the lowest being visits from outside the ZCTA (median: 2.4).

### Predictors of COVID-19 Incidence Risk

#### Global Model

Using a relaxed alpha of 0.15, several variables had potential univariable associations with COVID-19 incidence risk (**Table 2**). These included: (a) socioeconomic and demographic variables: percentage of male and Black populations, average household size and median household income; (b) all education and occupation variables, with the exception of the percentage of the population with an associate’s degree and those employed in retail and transportation; (c) all chronic disease and movement variables (except risks of hospitalizations attributable to tobacco use and COPD, within ZCTA visits and between ZCTA visits) (**Table 2**).

**Table 2:** Univariable associations between ZCTA-level COVID-19 risk and potential predictors in the Greater St. Louis Area, Missouri (USA)

| Type of Variable | Variable | Coefficient | 95% Confidence Interval | p-values |
| --- | --- | --- | --- | --- |
| **Demographic Factors** |  |  |  |  |
|  | % male population | 0.014 | -0.002, 0.030 | 0.110 |
|  | % black population | 0.003 | 0.001, 0.005 | 0.007 |
|  | % Hispanic/Latino population | 0.020 | -0.009, 0.050 | 0.168 |
|  | % over 65 | -0.007 | -0.019, 0.005 | 0.245 |
|  | Average household size | -0.150 | -0.330, 0.030 | 0.114 |
| **Educational Variables** |  |  |  |  |
|  | % with ≤ high school education | -0.003 | -0.007, 0.001 | 0.147 |
|  | % with some college education | -0.010 | -0.022, 0.001 | 0.079 |
|  | % with associate's degree | -0.007 | -0.028, 0.013 | 0.482 |
|  | % with bachelor's degree | 0.007 | 0.001, 0.013 | 0.024 |
| **Economic Variables** |  |  |  |  |
|  | % below poverty level | 0.002 | -0.004, 0.008 | 0.595 |
| | median household income (\$10,000s) | 0.017 | -0.002, 0.037 | 0.091 |
| **Occupation Variables** |  |  |  |  |
|  | % in agriculture<sup>1</sup> | -0.055 | -0.10, -0.005 | 0.030 |
|  | % in construction | -0.035 | -0.048, -0.021 | <0.0001 |
|  | % in retail | 0.001 | -0.010, 0.030 | 0.308 |
|  | % in transportation | 0.009 | -0.015, 0.033 | 0.478 |
|  | % in manufacturing | -0.019 | -0.032, -0.006 | 0.003 |
|  | % in education or healthcare | 0.011 | 0.001, 0.022 | 0.032 |
| **Health Behavior (ZCTA-level number of hospitalized patients that use tobacco per 100 persons)** |  |  |  |  |
|  | % tobacco<sup>2</sup> | 0.007 | -0.004, 0.018 | 0.245 |
| **Co-morbidities (ZCTA-level number of hospitalized patients with specific condition per 100 persons)** |  |  |  |  |
|  | % obesity | 0.047 | 0.017, 0.078 | 0.004 |
|  | % cancer | 0.059 | 0.024, 0.094 | 0.002 |
|  | % COPD<sup>3</sup> | 0.009 | -0.036, 0.053 | 0.692 |
|  | % CKD<sup>4</sup> | 0.046 | 0.021, 0.072 | 0.001 |
|  | % heart failure | 0.080 | 0.022, 0.140 | 0.007 |
|  | % diabetes | 0.035 | 0.010, 0.060 | 0.006 |
| **Movement variables** |  |  |  |  |
|  | Outside visits (1,000,000s) | 0.980 | -0.099, 2.100 | 0.074 |
|  | Within visits (1,000,000s) | 0.082 | -0.120, 0.290 | 0.427 |
|  | Between visits (1,000,000s) | 0.230 | -0.110, 0.570 | 0.171 |
|  | Per capita outside visits | 0.016 | 0.005, 0.028 | 0.004 |
|  | Per capita within visits | 0.008 | 0.003, 0.014 | 0.008 |
|  | Per capita between visits | 0.016 | 0.010, 0.022 | <0.0001 |

<sup>1</sup>Percent of population employed in agriculture, forestry, fishery and mining
<sup>2</sup>ZCTA-level % hospitalized patients that were tobacco users
<sup>3</sup>Chronic Obstructive Pulmonary Disease
<sup>4</sup>Chronic Kidney Disease

Based on the final global multivariable negative binomial model, the incidence risk of COVID-19 tended to be higher in ZCTAs that had high percentages of the population with bachelor’s degrees (p<0.0001) and those hospitalized due to obesity (p<0.0001) (**Table 3**). In contrast, COVID-19 risks tended to be lower in ZCTAs which had high percentages of the population employed in agriculture if the ZCTAs had zero per capita between ZCTA visits (**Table 3**). The significant (p<0.0001) interaction between percentage of the population in agriculture and the per capita between ZCTA visits implies that the latter was a significant effect modifier of the relationship between COVID-19 risk and percentage of the population employed in agriculture. Thus, although COVID-19 risk tended to be lower in ZCTAs that had a high percentage of the population employed in agriculture but no between ZCTA per capita visits, the risk of COVID-19 was higher in ZCTAs that had both high percentage of the population employed in agriculture and high per capita between ZCTA visits.

**Table 3:**
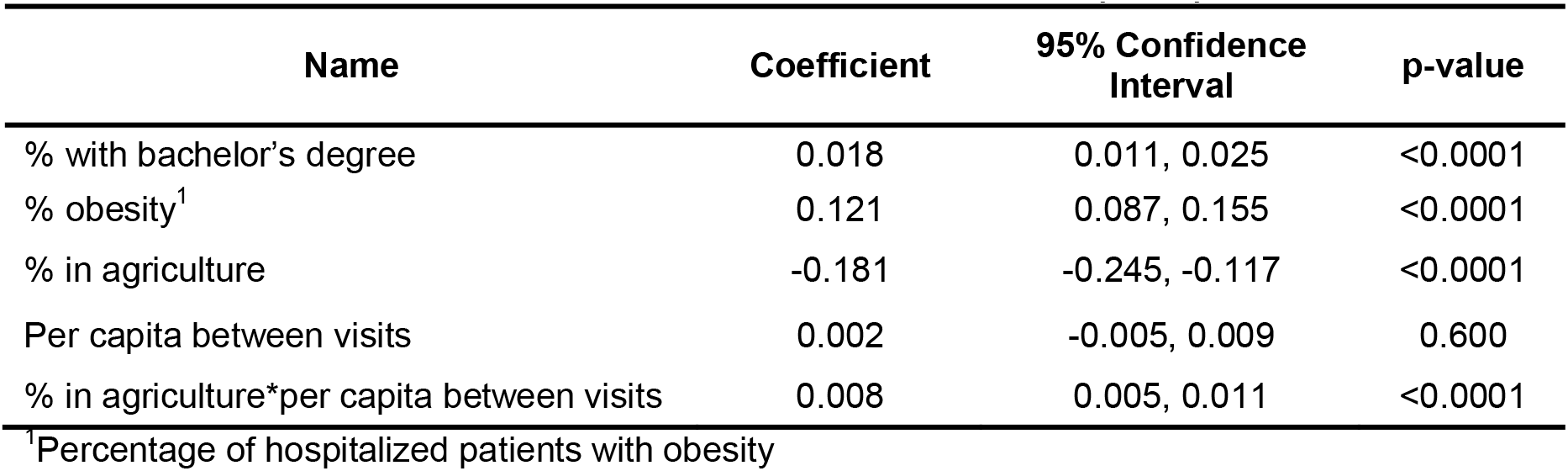
Final global negative binomial model showing significant predictors of COVID-19 risk in the Greater St. Louis Area, Missouri (USA)

The geographic distribution of COVID-19 risks and each of the predictors in the final model are shown in **Figure 2**. Note that the white patches in the maps indicate ZCTAs with missing data. The risks of COVID-19 were higher in the Eastern region of the study area, with pockets of high risks in ZCTAs located in St. Louis, St. Louis City and St. Charles counties while the lowest risks were observed in ZCTAs in Warren, Jefferson and Franklin counties. The distribution of the percentage of the population with bachelor’s degrees followed similar spatial patterns. The ZCTAs that had high percentages of the population employed in agriculture tended to have low COVID-19 risks (**Figure 2**).

**Figure 2.**
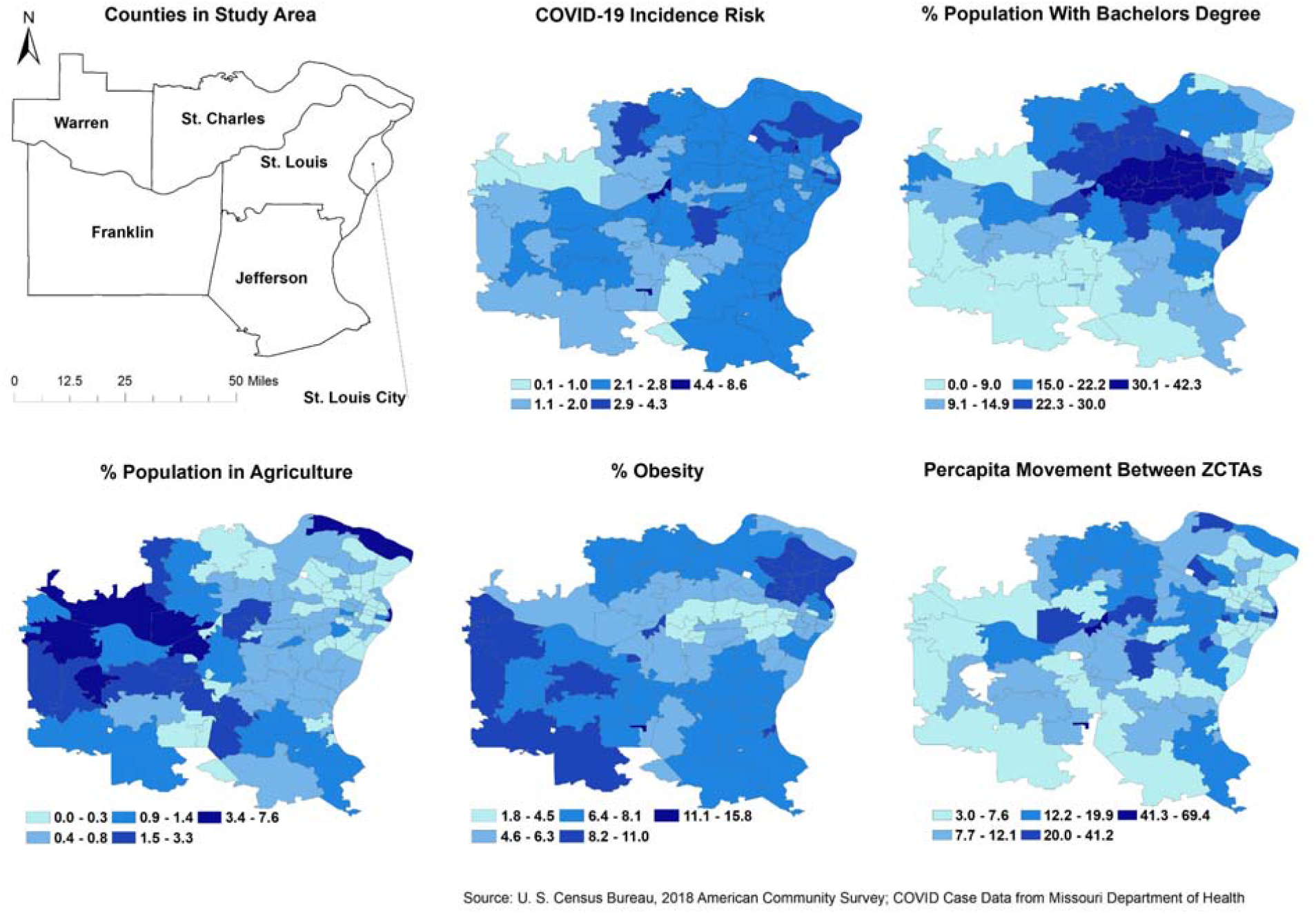
Geographic distribution of ZCTA-level COVID-19 risk and its significant predicators in the Greater St. Louis area, Missouri (USA).

#### Local Model

The randomization non-stationarity test results indicate that the coefficients of the association between percentage of the population employed in agriculture and COVID-19 risk was non-stationary (p=0.049) and, therefore, varied by geographic location (**Table 4**). This is further supported by the fact that the interquartile range of the GWNB model coefficient for percentage of the population employed in agriculture (0.1232) is greater than twice the standard error of the global coefficient (0.0652) for percentage of the population employed in agriculture (**Table 4**).

**Table 4:** Results of assessment of stationarity of the coefficients of the predictors of the COVID-19 risks in the Greater St. Louis Area, Missouri.

|  | NB SE<sup>1</sup> | NBSEx2 | GWNB<br>IQR<sup>2</sup> | IQR-2(SE) | GWNB<br>p-value | Is Coefficient<br>Non-Stationary? |
| --- | --- | --- | --- | --- | --- | --- |
| % with bachelor's degree | 0.0035 | 0.007 | 0.0067 | -0.003 | 0.102 | No |
| % in agriculture | 0.0326 | 0.0652 | 0.1232 | 0.058 | 0.049 | Yes<sup>4</sup> |
| % obesity<sup>3</sup> | 0.0174 | 0.0348 | 0.0255 | -0.0093 | 0.322 | No |
| Per capita between visits | 0.0035 | 0.0007 | 0.0034 | -0.0036 | 0.544 | No |
| % in agriculture*per capita<br>between visits | 0.005 | 0.01 | 0.0043 | -0.0057 | 0.085 | No |

<sup>1</sup>Standard Error from the Negative binomial model
<sup>2</sup>Interquartile range of the geographically weighted negative binomial standard error
<sup>3</sup>Percentage of hospitalized patients with obesity
<sup>4</sup>Coefficients are non-stationary based on both the p-value of the stationarity test and IQR-2(SE) assessment

The strength of the negative association between percentage of the population in agriculture and COVID risk varied considerably by geographic location and showed a gradient increasing from East to West, with the strongest association occurring in the West (**Figure 3**) which also has the highest percentage of the population employed in agriculture (**Figure 2**). When there is no (zero) per capita between ZCTA visits, the direction of this association stayed consistently negative across all ZCTAs in the study area (**Figure 3**). However, this association was modified and reversed by the number of per capita between ZCTA visits with COVID-19 risk increasing as both the percentage of population in agriculture and the number of per capita between ZCTA visits increased as evidenced by the significant interaction term (**Table 3**). It is worth noting that the coefficient of the interaction term was stationary (**Table 4**) implying that the strength of the modified association was the same across all ZCTAs in the study area.

**Figure 3.**
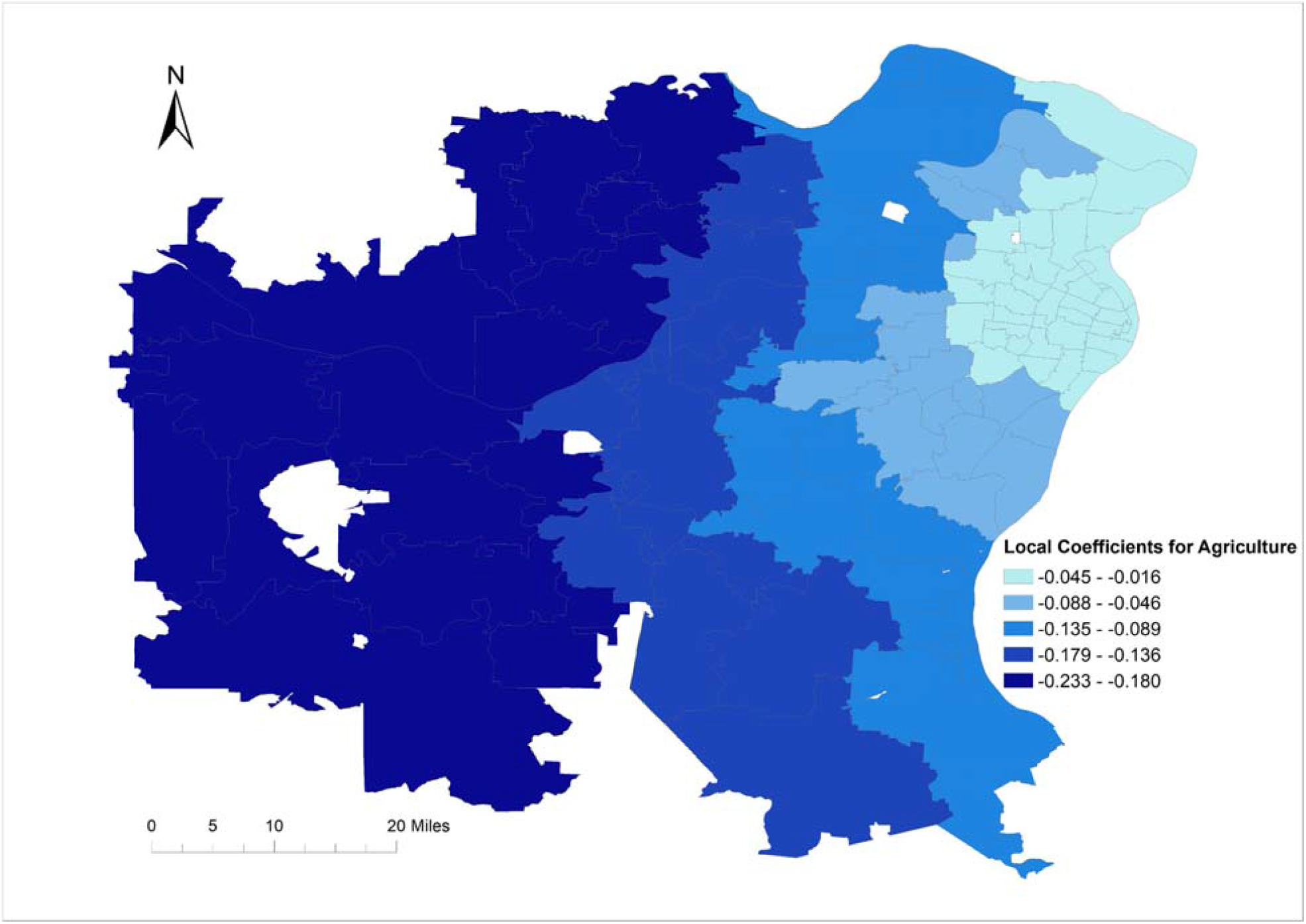
Geographically varying coefficients of local geographically weighted negative binomial model of COVID-19 risks in the Greater St. Louis Area, Missouri (USA)

## Discussion

This study identified ZCTA-level geographic disparities of COVID-19 risk in the greater St. Louis area of Missouri. It also investigated predictors of the identified disparities. The identified significant association between COVID-19 incidence risk and percentage of the population with bachelor’s degree is in contrast to findings from individual level studies that reported higher COVID-19 morbidity and mortality among individuals with lower educational attainment [25,26]. It also contrasts with reports of a study in a county in Michigan that reported no association between COVID-19 incidence risk and level of education [4]. This may suggest that the importance of specific predictors might vary by geographic location and might be driven by contextual factors. Additionally, the observed association between higher education attainment and COVID-19 risk observed in this study might be more a reflection of access to COVID-19 testing and not necessarily differences in disease incidence. It is likely that individuals with higher education attainment had better access to COVID-19 testing, than those with lower education attainment, especially during the time covered by the study when testing was not yet widely available.

The significant association between risk of COVID-19 and obesity hospitalization risks observed in this study is consistent with reports from several studies [27–30]. A number of reports have also linked obesity to more severe COVID-19 illness, hospitalizations and deaths [31–36]. A UK study reported that, compared to individuals of normal weight, overweight and obese individuals had 44% and 97% higher risk of COVID-19, respectively [37]. A pooled analysis conducted as part of a meta-analysis by Popkin *et al* reported that the odds of individuals with obesity testing positive for COVID-19 were 46% higher than among those who were not obese [36]. Some authors have suggested that excess ectopic fat deposition that occurs in obese individuals reduces protective cardiorespiratory reserve and potentiates the immune dysregulation that might mediate the progression to critical illness and organ failure in some COVID-19 patients [31].

Thus, obesity has harmful effects on lung function, reducing both forced expiratory volume and vital capacity [31]. Furthermore, obesity has modulatory effects on key immune cell populations critical in the response to SARS-CoV-2 virus. Increased anti-inflammatory cells inhibit the ability to reduce the infection, because inflammatory responses are needed to control viral spread [36]. This likely impairs the immune response to SARS-CoV-2. Obesity has also been reported to impair the development of immunological memory. For example, influenza vaccination in adults with and without obesity results in equivalent influenza-specific antibody titers at 30 days post vaccination, but antibody titers wane more among obese compared to non-obese adults 1 year after vaccination [36,38].

The inflammatory factors involved in obesity play a role in the development of severe lung diseases. There is evidence that susceptibility to acute respiratory distress syndrome, associated with COVID-19, is significantly greater among obese than non-obese individuals [39]. Moreover, obesity has been shown to increase the risk of influenza morbidity and mortality [40] due to impaired innate and adaptive immune responses [41]. It is suspected that COVID-19 vaccines might be less effective among individuals with obesity due to this weakened immune response [36]. These immunological impairments among obese individuals might suggest faster waning of immunity following vaccination among obese than non-obese segments of the population. This is particularly concerning considering the growing prevalence of obesity. Future individual level studies will need to focus on improving our understanding of the relationship between obesity and COVID-19 vaccination efficacy and waning of immunity to better guide vaccination regimens.

The COVID-19 pandemic has occurred at a time when the prevalence of obesity is increasing throughout the world [36] with almost all countries today having prevalence of overweight/obesity greater than 20% [42–44]. Unfortunately, the movement and social distancing restrictions due to the COVID-19 pandemic have resulted in changes in food consumption and physical activity patterns that might exacerbate current trends in the prevalence of obesity. It is concerning that these changes might have implications lasting way past the pandemic period [36] not only in the study area, but other geographic regions as well.

Although, in the absence of per capita between ZCTA visits, the percentage of the population engaged in agriculture had a negative association with COVID-19 risk, this association was modified and even reversed by per capita between ZCTA visits since the interaction term between the percentage of the population in agriculture and per capita between ZCTA visits was positive and statistically significant. Other studies have reported higher prevalence of COVID-19 in regions with high percentages of individuals engaged in agricultural activities [6,8]. Based on the findings of the current study, while the number of per capita between ZCTA visits did not have a significant association with COVID-19 risk on its own, it was an effect modifier of the effect of agricultural occupation on COVID-19 risk. This suggests that, in the Greater St Louis Area, per capita movements between ZCTAs may have a more profound impact on COVID-19 risk in the more rural communities (which have more individuals engaged in agriculture) than in urban locales.

It is worth noting that although both total visits (not adjusted for population) and per capita visits (adjusted for population) were assessed for potential association with COVID-19 risk, only the per capita visits had significant association with COVID risk. This may be an indication of the fact that it is not just mobility that is important, but that where the individuals go and the kind of interactions they have may be equally, if not more important. The lack of association between race and COVID risk is contrary to findings from other studies that have reported significant associations between Black population and COVID-19 risk [4,5,45].

The fact that the coefficients for the percentage of the population in agriculture were non-stationary and hence varied by geographic location implies that the importance and potential impact of this variable is not constant across the study area but varies based on geographic location. Thus, the strength of association between percent of population in agriculture and COVID-19 risk changes by location implying that a global coefficient was not sufficient in describing this relationship. Therefore, the focus of control efforts might need to vary based on location since some predictors may be more important in some locales than others.

### Strengths and limitations

There is currently lack of standardized data collection approaches across ZCTAs potentially resulting in differences in case ascertainment across ZCTAs. Moreover, hospitalization data can be influenced by differences in access to healthcare and may not always be representative of the population at large. A limitation of the safe SafeGraph data is that it may not be representative of all the segments of the population. For instance, it uses data collected by applications on only smartphones and thus does not include data on individuals who do not use smartphones. Some smartphone users may also opt-out from the service that collects mobility data and hence their data may not be collected. Additionally, for privacy reasons, SafeGraph data from ZCTAs with less than 5 devices were not included in the analysis. Therefore, SafeGraph data may not be representative of the true population mobility and hence the findings should be interpreted with this limitation in mind. The primary strength of this study is that it used both global and local models to identify predictors of COVID-19 risk. This approach allows identification of geographically varying regression coefficients and better descriptions of the associations between predictors and outcomes so as to better guide geographically targeted local control and prevention efforts. Another strength of the study is the use of SafeGraph population mobility data. The relevance of population movements to the COVID-19 infection risk is evident from the stay-at-home and social distancing directives that were put in place during the pandemic. The statistical analysis conducted in this study provided an insight into the possible associations between COVID-19 risk and various types of movements into and within ZCTAs.

### Conclusions

Geographic disparities of COVID-19 risks exist in the greater St Louis area. These disparities are, in part, explained by socioeconomic factors, chronic disease risks and population movement patterns that may be important in guiding efforts to control the disease and reduce disparities. Use of GIS, global, and local models help improve our understanding of geographic disparities in disease risks and the determinants thereof. The identified association between COVID-19 risk and obesity as well as the known immunological impairments among obese individuals is concerning considering the growing prevalence of obesity. Future studies will need to focus on improving our understanding of the relationship between obesity and COVID-19 vaccination efficacy and waning of immunity to better guide vaccination regimens.

## Data Availability

The investigators cannot share the datasets, which are not publicly available, because they do not have legal ownership nor authority to share the data. However, the data can be requested from the Missouri Hospital Association at 4712 Country Club Dr, Jefferson City, MO 65109, telephone (573) 893-3700.

## Declarations

### Ethical approval and consent to participate

This study was approved by the North Carolina State University Institutional Review Board (IRB number: 22342) and all study methods were carried out in accordance with relevant guidelines and regulations. The study used anonymized secondary data provided to the investigators, by the Hospital Industry Data Institute, in such a manner that the identity of human subjects cannot be ascertained directly or through identifiers linked to the subjects. The investigators did not contact the subjects and did not re-identify subjects.

### Consent for publication

Not applicable.

### Authors’ contributions

SL, CL, ALL, and AO conceptualized research idea

LL collected and curated the data

PD, MI, SL, CL, ALL, and AO analyzed data

PD, MI, and AO wrote the manuscript

All authors edited the manuscript

All authors read and approved the final manuscript.

## Acknowledgements

We thank the St. Louis Comparative Modeling Network for facilitating access to data and useful discussions.

